# The Hidden Curriculum of Autism Care: Informal Learning among Clinicians and Caregivers in Kazakhstan

**DOI:** 10.1101/2025.05.27.25328431

**Authors:** Faye Foster, Akbota Kanderzhanova, Akbota Tolegenova, Paolo Colet, Valentina Stolyarova, J Gareth Noble

## Abstract

**Background:** Autism care in Kazakhstan continues to develop amid limited training, unclear referral systems, and uneven provider readiness. While gaps in formal infrastructure have been well documented, the informal adaptations used by families and Physicians have received less attention.

**Aim:** To explore how informal learning, peer knowledge, and role modelling shape autism diagnosis and care in Kazakhstan, by examining the everyday behaviours and improvised routines that form a hidden curriculum alongside formal systems.

**Methods:** This study uses secondary analysis of two datasets: semi-structured interviews with parents of autistic children (n = 10), and national survey data from parents (n = 190) and physicians (n = 110). A concept-driven thematic analysis was applied to both interview and survey responses to identify recurring informal behaviours and decision-making patterns. Descriptive survey statistics were used to contextualise these qualitative findings.

**Results:** Parents frequently reported being left without post-diagnostic guidance, prompting them to rely on social media, peer groups, and personal networks. Physicians, especially those early in their careers, described relying on observation or mimicking more experienced colleagues in the absence of formal training. These behaviours reflect a hidden curriculum, unofficial but influential practices that shape practices that lead to inconsistent post-diagnostic guidance.

**Conclusion:** In Kazakhstan, informal knowledge-sharing and improvisation often affect care consistency or outcomes. These practices help fill urgent gaps but also introduce inconsistencies and shift burdens onto families, especially mothers. Policy and training reforms must acknowledge and address these informal structures to improve equitable care.

## Introduction

In many low- and middle-income countries (LMICs), including Kazakhstan, autism care systems are developing in the context of persistent challenges such as limited training, unclear diagnostic pathways, and weak institutional support. In Kazakhstan, “weak institutional support” often means that families are given a diagnosis without follow-up services, there are few autism-specific specialists, and coordination between healthcare, education, and social services is minimal or inconsistent. As a result, families are often left to navigate care on their own (An et al., 2018, 2020; Ghanouni & Naimpally, 2025; Pervin et al., 2022). While these issues are well-documented, less attention has been paid to how families and professionals adapt in practice, through informal routines, imitation, and shared knowledge (Behmanesh et al., 2025; Silveira et al., 2024; Zhang et al., 2025).

Drawing on interviews and national survey data from both parents and physicians in Kazakhstan (Foster et al., 2025a & 2025b), this study examines the unspoken behaviours that shape autism care in Kazakhstan, such as physicians making a diagnosis after only a few minutes of observation, and parents turning to Telegram chats or Instagram for guidance in the absence of formal support. For instance, junior physicians imitating the practices of their more experienced colleagues because standardized protocols were missing, while mothers often rely on social media, peer networks, and informal contacts to guide their care decisions. These behaviours reflect what some researchers call a ‘hidden curriculum’, norms and knowledge passed on through experience rather than formal teaching (Hafferty, 1998; Lawrence et al., 2018). Similarly, the concept of lay expertise helps explain how parents learn to spot signs, choose therapies, and make care decisions on their own, often because no one in the system is telling them what to do (Eyal & Medvetz, 2023).

Kazakhstan shares many of the same challenges found in other LMICs, including delayed diagnoses, limited access to specialists, and widespread use of complementary and alternative medicine (CAM), often out of necessity rather than preference (An et al., 2018, 2020). Research on provider readiness in Kazakhstan also points to inconsistent training and low confidence among physicians, especially in mainstream settings (Kosherbayeva et al., 2024a; 2024b). However, these studies tend to focus on formal knowledge and do not examine the informal ways in which people learn to provide care.

This study builds on earlier research by re-examining interviews and survey responses through the lens of informal learning, mimicry, and peer support. Its goal is to identify the hidden curriculum that shapes care in a system where formal guidance is often lacking.

## Aim

The aim of this study is to explore how informal learning processes shape autism diagnosis and care in Kazakhstan. Through secondary analysis of qualitative interviews and two national surveys, it examines how caregivers and professionals learn to act in the absence of structured training. By highlighting these informal practices, the study contributes to a better understanding of how care is delivered in low-resource settings.

## Methods

### Study Design and Rationale

This paper draws on secondary analysis of two previously conducted studies on autism care in Kazakhstan, carried out in 2024. While the original research focused on diagnostic experiences, healthcare communication, and service access, both datasets contained repeated references to informal learning, improvisation, and social transmission of knowledge. This analysis specifically examines those aspects, applying the lens of the “hidden curriculum” to make sense of how clinical practice and parental knowledge are shaped not through formal instruction, but through daily interactions, trial-and-error, and peer-to-peer advice.

The hidden curriculum here refers to the unspoken norms, behaviours, and habits that influence how autism care is delivered and received, ranging from how diagnoses are made to how families learn to navigate the system. The study focuses on those points where official guidance was lacking or unclear, and where families and frontline Physicians appeared to fall back on tacit rules, social mimicry, or community-driven knowledge.

### Data Sources

This manuscript draws on data from two preprint studies conducted by the authors in Kazakhstan. Together, these studies offer a multi-perspective view of informal learning, diagnostic challenges, and adaptive caregiving within Kazakhstan’s autism care system.

**Study 1:** This study used qualitative interviews with parents of autistic children to better understand their experiences with the healthcare system in Kazakhstan. Ten participants were recruited using a mix of social media advertisements, contact follow-up from an earlier survey, and referrals from other parents (snowball sampling). Families came from both rural and urban settings.

Interviews took place between January and April 2024. They were held online to make participation easier across Kazakhstan’s large geographic area. Interviews were conducted in Kazakh or Russian, transcribed, then translated into English. A second translator reviewed the translations to check for accuracy.

The interviews followed a semi-structured guide that focused on early interactions with healthcare professionals, the diagnostic process, follow-up care, and challenges faced. Interview recordings lasted 60–90 minutes. Data saturation, when no new themes emerged, was reached after ten interviews.

Ethical approval was granted by Nazarbayev University School of Medicine (Ref: 2024Feb#02), and informed consent was obtained from all participants. This study is available as a preprint (Foster et al., 2025a).

**Study 2:** This cross-sectional study used online surveys to compare how parents and physicians in Kazakhstan view the autism diagnosis and care process. Two separate questionnaires were created, one for each group, and shared between February and August 2024.

The parent survey had 190 respondents, mostly mothers, and focused on early signs of autism, diagnosis timelines, and what types of support were offered after diagnosis. The physician survey had 110 respondents, mainly psychiatrists, and asked about diagnostic practices, beliefs about autism, and training experiences. Both surveys were offered in Kazakh and Russian. Both surveys had national coverage, with responses from all major regions in Kazakhstan.

Parents were recruited through rehabilitation centres, support groups, and social media. Physicians were contacted through hospitals, email, and professional networks. Participation was voluntary, and all responses were anonymous. Ethical approval was granted by the Nazarbayev University School of Medicine (Ref: 2024Feb#04 and 2024Feb#05). This study is available as a preprint (Foster et al., 2025b).

### Data Analysis and Reflexivity

This study used a concept-driven thematic analysis to explore how informal learning shapes autism care in Kazakhstan. We drew on the concept of the hidden curriculum: unofficial, unspoken, and often unintended ways that knowledge, norms, and behaviours are passed on outside formal instruction. Building on prior research on informal clinical learning and lay expertise, we focused on how caregivers and providers adapt when institutional support is limited. These concepts shaped both how we selected the data and how we interpreted what participants shared.

The analysis focused on how participants, particularly parents, acquired knowledge, adapted to challenges, and made care decisions lacking professional guidance or structured support. For example, parent statements like “doing everything myself,” or physicians citing “intuition” rather than screening tools, were treated as evidence of informal learning. The goal was not to recode the entire dataset but to identify patterns that reflected how people navigated care in a fragmented system.

The lead researcher read all transcripts multiple times to develop initial codes. Two additional team members reviewed and refined the coding framework on a subset of transcripts. Discrepancies were discussed in team meetings, and the codebook was revised accordingly. Related codes were grouped into broader categories and eventually organized into four final themes:

1. Parent expertise as informal curriculum
2. Absence of formal guidance and role confusion
3. Mimicry and improvisation in clinical behaviour
4. The emotional economy of informal knowledge

To ensure rigour, the team met regularly to reflect on coding decisions and challenge interpretive assumptions. Although this was a secondary analysis, it followed a systematic and transparent process. The team brought a range of clinical, academic, and cultural perspectives, including direct experience in autism care, qualitative research, and Kazakhstani healthcare systems. These perspectives shaped how we read the data and prompted ongoing reflection on our own positionality.

Survey results were reviewed after the thematic coding process and used to contextualize the qualitative findings. Descriptive statistics from both parent and physician surveys provided additional insight by highlighting areas of convergence or contrast with the interview data. These included parent-reported clarity of post-diagnosis pathways, access to autism-related information, financial burden, and physician-reported use of screening tools and referral protocols. The quantitative data were not re-analysed for statistical significance but served to frame and support the interpretation of the qualitative themes.

## Results

This section presents four interlinked themes that reveal how informal learning, social mimicry, and parental adaptation shape autism care delivery in Kazakhstan. Drawing on interviews with parents and supported by national physician survey data, the analysis traces how informal knowledge becomes a substitute for structured support, producing a “hidden curriculum” that guides both families and frontline providers.

### 1. Parent Expertise as Informal Curriculum

In the absence of structured guidance, parents became informal educators, passing on practical strategies for navigating care systems and managing daily challenges, both for themselves and others. This transfer of knowledge reflects a hidden curriculum in which parental expertise, rather than professional instruction, becomes the foundation for navigating autism care.

Several interviewees described turning to social media as a first step in making sense of the diagnosis. For example: “I started searching Instagram and following mom bloggers with the same diagnosis.” (Parent 3). This was more than emotional curiosity, it was a deliberate act of self-training, prompted by the absence of reliable information from providers.

Other parents reflected on the quality and credibility of peer knowledge: “The mothers in the Telegram groups were more informed than some neurologists.” (Parent 1). Such comments suggest that parents were not only filling a gap left by the health system but also challenging its authority.

Informal parent groups on WhatsApp and Telegram became spaces of exchange, where information was circulated, tested, and adapted: “In all these chats, we share information with each other.” (Parent 7) and “MamLife, WhatsApp chats… I read what I need there.” (Parent 2).

Parents were also self-organising across language and platform boundaries, drawing on multilingual resources to build an evidence base when local materials were unavailable: “I searched articles in English if I couldn’t find them in Russian. Googled, read, watched.” (Parent 3).

Others gained access to these communities through peer referral, highlighting the relational fabric of informal expertise: “A mom whose son had autism gave me all the links. I didn’t even know these groups existed.” (Parent 5).

These networks served as a form of grassroots infrastructure, providing both emotional affirmation and practical guidance. In this way, informal parent knowledge became an improvised curriculum; a growing, family-to-family network where advice was passed along, adapted through experience, and shaped by the specific challenges each family faced.

Survey data underscore the institutional gap that gave rise to this knowledge-making. Only 32% of parents reported receiving informational materials at the time of diagnosis, and fewer than 25% were referred to any parent training or support services. This vacuum of formal guidance left families no choice but to turn to one another.

### 2. Absence of Formal Guidance and Role Confusion

Without clear post-diagnostic pathways, both caregivers and clinicians are left uncertain about their roles. This lack of structure creates a hidden curriculum in which families are left to make up their own next steps, tracking down therapies, coordinating appointments, and deciding what to do next, all tasks that often fall to mothers by default. One parent recalled the abruptness of the encounter: “No one clearly explained what to do after the diagnosis. They just said, autism, and that was it.” (Parent 3). Another parent recalled being referred vaguely without a plan: “We were just left on our own.” (Parent 4)

Even when care was sought proactively, parents were passed between providers with no clear responsibility: “They just bounced me around, neurology, then back again, and said they couldn’t help. I was told to see a local neurologist… and to come back when he’s four.” (Parent 1).

For some, the lack of advice became a point of lasting frustration: “Why didn’t she advise us? Why didn’t she say to get started… I would’ve never known.” (Parent 5).

Parents frequently described situations in which they could identify problems, but professionals failed to act or guide them: “I saw the symptoms… but found no support from physicians. So, parents just don’t know where to go next.” (Parent 9).

This absence of clear pathways also affected physicians, who appeared uncertain themselves. One mother observed: “They [the physicians] seemed just as confused as we were.” (Parent 5). Another described a provider recommending institutionalisation, not treatment: “Physicians say everything’s fine. I went to psychiatry, they said, ‘just hospitalize him.’ I thought, what are we supposed to do?” (Parent 9).

Survey findings support these experiences. 70% of parents reported being unclear on what steps to take after diagnosis, while 56% of physicians acknowledged the lack of a standardised referral protocol adding to the reported lack of training in autism. Although there is an official standard medical protocol for autism in Kazakhstan as of 2021, where ADI-R, ADOS-2, and M-CHAT-R are recommended as gold standards for autism diagnostics and screening, the guidelines are currently based on ICD-10 and might need ICD-11 based careful revisions. Along with the affordable access to Russian/Kazakh diagnostic instruments across all the regions of the vast country, state-of-the-art validated Kazakh adaptations of the screening and diagnostic tools might also be required to address the needs of Kazakh speaking citizens. This dual uncertainty points not just to poor communication, but to a system in which no clear referral or follow-up process exists, leaving families to figure out next steps entirely on their own.

### 3. Mimicry and Improvisation in Clinical Behaviour

With limited formal autism training, many healthcare providers learn by copying how colleagues deliver diagnoses or by making judgments based on brief observations rather than using standardized screening tools. This informal modelling process, largely invisible and unacknowledged, embodies the hidden curriculum at work in clinical settings.

Parents often described receiving diagnoses based on brief observation or unstructured impressions, rather than formal screening tools: “They gave us the diagnosis just by looking.” (Parent 6). These experiences suggest a diagnostic culture shaped more by tacit norms and observational habits than clinical training.

In other cases, junior staff appeared to replicate the actions of their seniors: “The younger physician repeated exactly what the senior one said, without even examining the child.” (Parent 7). Rather than deliberate malpractice, this reflects a process where junior staff replicate the diagnostic methods of senior colleagues rather than follow established guidelines, because structured mentorship or training is often missing.

One parent described a diagnosis given after mere observation: “The physician said, ‘Calm down, mom, I want to observe his behaviour’… just watched the child… then said, ‘your child has autism.’” (Parent 3).

Others recounted highly improvised diagnostic procedures: “I came in, and the physician just said he’s too young. Later, at PMPK, they just told me to hold him still. I did, and after 15 minutes they handed me a referral.” (Parent 5).

Survey data reinforce this pattern: while fewer than 30% of physicians reported using tools like M-CHAT, nearly 90% said they felt confident in identifying autism. This suggests that confidence is being shaped more by habit and professional hierarchy than by formal instruction. In effect, this creates an informal learning system where routines are passed down and rarely questioned, leading to inconsistency in how autism is recognized and diagnosed.

### 4. The Emotional Economy of Informal Knowledge

Trying to navigate care without clear guidance took a toll on families, resulting in stress, financial strain, and emotional exhaustion, especially for mothers, who were left to manage nearly every aspect of care alone. Over time, they learned to manage their emotions quietly and persist through uncertainty, because the system offered little support in moments of crisis. In this way, emotional resilience became part of the hidden curriculum, something mothers were expected to develop without being shown how. The sense of urgency created by unclear guidance and long wait times often led to sacrifices made in isolation: “We sold our car to pay for therapy. I felt I couldn’t wait for the system to figure it out.” (Parent 10) Caring duties also fractured daily life: “Your personal life disappears. You can’t work; you can’t trust anyone to watch him, not even family.” (Parent 1)

Some described making financial decisions under duress: “When you’re in a state of panic, you think you have to take everything offered… Parents go into debt.” (Parent 3).

Moments of acute stress were often compounded by the lack of professional support during crises:

> “When I saw it, I was terrified… He was running around the room and banging his head against the wall. It was terrifying. I cried and called the ambulance.” (Parent 1) “It’s very frightening… because panic starts inside you, and you just run.” (Parent 3) “I was scared… he was hitting his head against the wall… it was unbearable to watch.” (Parent 7)

These accounts reveal the impact of a fragmented system: delayed diagnoses, long waits for services, and constant uncertainty that leaves parents anxious, isolated, and financially overextended.

Survey findings mirror these struggles: 62% of parents reported paying out of pocket for therapies, and only 18% said they felt supported by the healthcare system after diagnosis. The emotional labour and financial risk involved in navigating autism care remain largely unacknowledged by existing support structures.

## Discussion

This study examined the informal behaviours, routines, and sources of knowledge that shape autism care in Kazakhstan. Rather than focusing solely on training deficits or service gaps, it explored how parents and physicians navigate care through unofficial, often improvised practices. These include relying on intuition or imitation in diagnosis, using peer networks for information, and managing care decisions where structured guidance is missing. Together, these practices form what has been described as a “hidden curriculum”, a set of unspoken norms and adaptations that operate alongside, and at times in contradiction to, formal policy and training structures (Hafferty, 1998). Adapting the current medical clinical protocols for autism to ICD-11 diagnostic criteria may both be challenging yet necessary to advance and enhance the diagnostic procedures and evidence-based care practices to adhere to the best international standards of autism management (Simashkova et al., 2019).

### Informal Knowledge as a Response to Institutional Silence

One of the clearest patterns in the data was the extent to which parents turned to informal learning to guide care. This included WhatsApp and Telegram groups, Instagram accounts of other mothers, and translated online articles. These networks offered practical advice, emotional support, and a form of validation that many parents did not find in clinical settings. Rather than passive recipients of care, parents became active agents in making sense of their child’s condition, taking on the role of case managers, educators, and advocates. Interestingly, rather than framing this as empowerment, parents often saw it as something they were forced to do because no one else was guiding them. This mirrors findings in other low- and middle-income countries where parents become informal care coordinators out of necessity, not choice (An et al., 2018; Kosherbayeva et al., 2024). A recent study conducted in Russia also found a lack of autism knowledge and expertise among healthcare professionals and indicated that parents of autistic children play a major role and initiative in noticing and detecting autism in their children (Mukhamedshina et al., 2022). This mirrors findings in other low- and middle-income countries where parents become informal care coordinators out of necessity, not choice (An et al., 2018; Kosherbayeva et al., 2024).

This informal knowledge, developed through necessity, forms part of the hidden curriculum of autism care. It’s not taught, not paid for, and rarely supported, yet it shapes how children access services and how families navigate a fragmented system.

Survey data reinforce this informal infrastructure. Only one-third of parents received any written information at diagnosis, and fewer than one in four were referred to support groups or training. In this context, informal peer knowledge is not supplementary, it is central.

### Learning to Diagnose Without Tools

Alongside parental adaptation, the findings point to an informal pattern of clinical learning, in which newer physicians appear to diagnose autism by watching more senior colleagues or relying on their own impressions. Parents reported that some Physicians made judgments after only brief observation, often without using screening tools. Survey data show that while fewer than 30% of physicians used validated instruments, nearly 90% reported feeling confident identifying autism. This gap suggests that confidence is being shaped by habits and role modelling, not formal training. In terms of “lay knowledge” concept, it is further argued that a majority of patients and their caregivers do not actually actively engage and participate in knowledge development, even though some individuals may engage in research studies or gain some specialized expertise (Wilcox, 2010).

This kind of learning works like an informal apprenticeship, where practical routines are learned informally and passed along without evaluation. In contexts where formal continuing education is weak and clinical oversight limited, such practices are understandable, but they can also produce inconsistency, delay, and misdiagnosis (Lee et al., 2022; Adams et al. 2021). These adaptations often reflect broader health system gaps and institutional uncertainty in resource-limited settings (Behmanesh et al., 2025; Silveira et al., 2024).

### The Costs of Navigating Alone

Across many interviews, parents, mostly mothers, spoke about the emotional weight of making major decisions without clear support. Some had to take drastic steps, like selling a car or leaving a job, just to pay for therapy. These were not long-term plans, but urgent responses to immediate needs. In moments of crisis, feelings of panic, isolation, and fear were common. Families often received a diagnosis but no clear next steps, leaving them to navigate the system on their own.

A recent Kazakhstani study found that most parents of autistic children do not feel supported after diagnosis. Stress, anxiety, and depression were widely reported 61%, 52.9%, and 53.7% respectively (Alibekova et al., 2022). Only 18% said they received enough support from the healthcare system. When no clear pathways were available, many turned to friends, other parents, or online groups for advice and reassurance.

This lack of formal guidance places much of the burden on mothers. Research from other low- and middle-income countries shows a similar pattern: women tend to carry most of the daily responsibilities for children with autism. The emotional toll, financial strain, and constant management demands are often overlooked when healthcare systems are designed (Mohammad et al., 2022; Salomone et al., 2023).

These pressures weren’t side effects; they were central to how families figured out what to do. Emotional resilience and self-control became survival tools, learned through experience rather than training. In that sense, emotional management becomes part of the hidden curriculum: something families pick up on their own because the system doesn’t show them how. These struggles don’t just reflect personal hardship; they show how uneven and unsupported the path can be for families trying to do the best for their children.

These struggles don’t just reflect personal hardship; they show how uneven and unsupported the path can be for families trying to do the best for their children.

In almost every family we spoke to, it was the mother who took on the responsibility of navigating care. Fathers were rarely present in interviews or surveys, which reflects everyday realities in Kazakhstan, where caregiving, especially for children with disabilities, is still largely considered a mother’s role and it also reflects how health and support systems are designed with the assumption that mothers will fill the gaps. (Alibekova et al., 2022; Mohammad et al., 2022; Salomone et al., 2023; Gallaher et al., 2023).

### Implications for Policy and Practice

These findings suggest that to improve autism care in Kazakhstan, policy efforts must engage not only with what professionals and families are formally taught, but also with how they learn to act in the absence of guidance. Informal practices emerge as coping strategies when formal systems fall short—but when left unaddressed, they can perpetuate inconsistency, delay, and inequity.

First, post-diagnosis pathways must be made more visible and consistent. This could include implementing a standardized follow-up checklist for physicians, automatic referrals to certified support services, and brief, language-accessible information packets for families. Evidence from initiatives like the HANDS in Autism® model and Anna Freud Centre’s programs suggests that such tools improve family engagement and service uptake (HANDS in Autism®, n.d.; Anna Freud Centre & AT-Autism, 2025).

Second, early-career physicians need more than guidelines—they need practical, supervised experience. In contexts like the HANDS in Autism® program in the U.S. and team-based mentorship models used in Kenya and India, case-based simulations and in-situ coaching have led to measurable increases in diagnostic confidence and earlier identification rates (Gallaher et al., 2023; Dhuga et al., 2022).

Third, parent networks shouldn’t be treated as a backup plan, they are already doing critical work. In the UK, the Anna Freud Centre has integrated peer-led groups into post-diagnostic support pathways, offering structured spaces for emotional support and shared learning (Anna Freud Centre & AT-Autism, 2025). Similar models could be adapted in Kazakhstan, with co-designed materials and modest funding to build trust and continuity. (Yi et al., 2020; O’Neill & O’Donnell, 2024).

Finally, national autism policy must address not only the technical training of health professionals, but also the emotional and logistical labour families, particularly mothers, perform to hold the system together. International research highlights that caregiving burdens are disproportionately shouldered by women, and that failure to address this leads to long-term emotional and financial strain (Mohammad et al., 2022; Salomone et al., 2023; Gallaher et al., 2023).

Understanding how people actually learn and act, what they improvise, mimic, or quietly absorb, should be the starting point for reforms. Only then can we begin to reduce diagnostic delays, ease the burden on families, and move toward a more consistent and equitable system of autism care.

## Limitations

This study used secondary analysis of existing qualitative interviews and survey data, which limited the ability to observe informal learning and clinical practice in real time. However, revisiting these data with a focused analytic lens allowed for deeper insight into how participants described their experiences and made sense of autism care without formal structures in place. Future research could benefit from ethnographic or longitudinal designs that capture how informal routines evolve, particularly as more training opportunities and tools become available in Kazakhstan.

While the sample included participants from different regions and care settings, it may not fully reflect the experiences of families in the most remote or underserved areas. Additionally, the voices of fathers were largely absent from both the interview and survey samples. Further work is needed to understand how paternal roles and broader family dynamics shape autism care, especially in contexts where caregiving responsibilities are heavily gendered.

## Conclusion

This study met its aim of exploring how informal learning, peer knowledge, and role modelling shape autism care in Kazakhstan. By analysing the experiences of both parents and physicians, we identified a hidden curriculum made up of intuitive decision-making, mimicry, and parent-driven knowledge networks that fill the gaps left by inconsistent training and unclear post-diagnosis pathways.

This study shows that in Kazakhstan, informal learning doesn’t just fill the gaps left by formal systems, it actively shapes how autism care is delivered, often working alongside or in place of official guidance. In the absence of consistent training or post-diagnosis guidance, both parents and physicians rely on intuition, imitation, and social networks to navigate uncertainty. These practices form a hidden curriculum that shapes how care is delivered, and decisions are made, often in ways that reproduce inequality and increase emotional and logistical burdens, especially for mothers.

These informal routines aren’t just gaps in the system; they’re how people cope with the system’s failure to provide clear post-diagnostic support, accessible services, and reliable guidance. However, relying on them without support risks reinforcing the very problems they are meant to solve. If future reforms aim to improve early diagnosis and equitable access to care, they must take these everyday realities seriously. This means addressing not only what physicians are taught, but how norms, habits, and knowledge are passed on through informal channels. Building more reliable, inclusive services will require bridging the gap between official systems and the ways people already learn and act within them.

## Data Availability

All data produced in the present study are available upon reasonable request to the authors

## Author Contributions

**Faye Foster:** Conceptualization, Methodology, Investigation, Writing – Original Draft, Supervision.

**Akbota Kanderzhanova:** Investigation, Writing – Review & Editing.

**Akbota Tolegenova:** Investigation, Writing – Review & Editing.

**Paolo Colet:** Writing – Review & Editing, Supervision.

**Valentina Stolyarova:** Writing – Review & Editing.

**J. Gareth Noble:** Conceptualization, Writing – Review & Editing, Supervision.

